# Large-Scale Longitudinal Comparison of Urine Cytological Classification Systems Reveals Potential Early Adoption of The Paris System Criteria

**DOI:** 10.1101/2022.05.06.22274783

**Authors:** Joshua J. Levy, Xiaoying Liu, Jonathan D. Marotti, Darcy A. Kerr, Edward J. Gutmann, Ryan E. Glass, Caroline P. Dodge, Louis J. Vaickus

## Abstract

Urine cytology is used to screen for urothelial carcinoma in patients with hematuria or risk factors (e.g., smoking, industrial dye exposure) and is an essential clinical triage and longitudinal monitoring tool for patients with known bladder cancer. However, urine cytology is semi-subjective and thus susceptible to issues including specimen quality, inter-observer variability, and “hedging” towards equivocal (“atypical”) diagnoses. These factors limit the predictive value of urine cytology and increase reliance on invasive procedures (cystoscopy). The Paris System for Reporting Urine Cytology (TPS) was formulated to provide more quantitative/reproducible endpoints with well-defined criteria for urothelial atypia. TPS is often compared to other assessment techniques to justify its adoption. TPS results in decreased use of the atypical category and better reproducibility. Previous manuscripts comparing diagnoses pre- and post-TPS lacked a longitudinal component and thus failed to consider temporal differences between diagnoses made under prior systems and TPS. By aggregating across time, studies may underestimate the magnitude of differences between assessment methods. We conducted a large-scale longitudinal reassessment of urine cytology using TPS criteria from specimens collected from 2008 to 2018, prior to the mid-2018 adoption of TPS at an academic medical center. Findings indicate that differences in atypical assignment were largest at the start of the period and these differences progressively decreased towards insignificance just prior to TPS implementation. This finding suggests that cytopathologists had begun to utilize the quantitative TPS criteria prior to official adoption, which may more broadly inform adoption strategies, communication, and understanding for evolving classification systems in cytology.

## Introduction

Urothelial carcinoma is a cancer that demands attention because it is both common (7th leading cause of malignancy) and has the highest recurrence rate of any cancer (70%) ^1,2^. Diagnostic protocols and clinical triage are based on a combination of urine cytology, cystoscopy +/-biopsy, and immunocytochemical and molecular studies, with resultant longitudinal follow-up for negative and atypical findings. While cystoscopy followed by biopsy with histological examination is the accepted standard for diagnosis, urine cytology is the preferred mode for detection and screening because: it is inexpensive; has a rapid turn-around time; and specimens are relatively easy to obtain. Abnormal cytology findings usually trigger more invasive and expensive assessment methods (e.g., cystoscopy, FISH) ^4,5^, though further investigation is important because these assessments determine treatment options, ranging from follow-up urine cytology to chemotherapy or surgery. This sequence is not entirely foolproof since abnormal cytological findings are subject to a high “false positive” rate (i.e., urine specimen is perceived to be abnormal while cystoscopy/biopsy leads to negative finding). Cytology-based examination is the currently accepted method for the detection of high-grade urothelial carcinoma (HGUC) while assessment of specimens for low-grade urothelial carcinoma (LGUC) is subject to variable cytomorphological features, which limits confidence in the findings. High inter-rater variability and equivocal findings are factors which still adversely impact urine cytology for screening ^6,7^. Surgical biopsy, while often considered the gold standard for bladder cancer is also fraught with sampling issues that may result in a negative biopsy on an unequivocally positive cytology result ^8^.

Several reporting and classification systems have been developed for assessing Urine Cytology (UC), yet many such systems failed to gain widespread adoption due to inconsistent terminology and grading criteria or overly complex scoring metrics ^9^. As assessment perspectives have shifted toward quantitative endpoints of cytological atypia, The Paris System for Reporting Urinary Cytology (TPS), conceived of in 2013 and officially published in 2016, has emerged as the widely agreed upon classification system for urine cytology bladder cancer screening ^2^. Cytologists using TPS typically assess the individual urothelial cells of a cytology specimen for features of atypia (nuclear hyperchromasia, high nuclear to cytoplasm area ratio (NC ratio), nuclear membrane irregularities, chromatin abnormalities), culminating in the assignment of UC diagnostic categories (four ordered classes: negative, atypical, suspicious, positive). Assignment to diagnostic categories typically leads to the following triaged responses: 1) interval follow-up if negative, 2) follow-up and potentially ancillary tests if atypical, and 3) cystoscopy and biopsy if suspicious or positive. Diagnosis of high-grade urothelial carcinoma (positive) includes following criteria: 1) at least 5 malignant urothelial cells, 2) NC ratio at or above 0.7, 3) markedly irregular nuclear membrane, 4) coarse/clumped chromatin and 5) nuclear hyperchromasia. A suspicious UC finding denotes uncertainty for a positive diagnosis but assignment into either class (suspicious/positive) often leads to the same disease management as aforementioned. At times, not all diagnostic criteria for malignancy may be satisfied due to a variety of confounding reasons (e.g., variable/confluent cellularity, unreliable NC ratio estimates, degenerated cells, specimen preparation), which may preclude ordering of a cystoscopy or biopsy. Hypothetically, the cystoscopy or biopsy could yield positive results if the tests were ordered ^10–15^. While high inter-observer variability and lower levels of diagnostic precision are still reported for TPS for some of the diagnostic categories, TPS is still preferred to the prior mode of cytological assessment (assigned acronym: PMC) ^16^.

Though TPS aimed to reduce this tendency, a remaining impediment to reliable urine cytology assessment is the assignment of “atypia”, which serves as a “catch-all” term often overused by pathologists to hedge against a negative, suspicious or positive UC diagnosis ^17–19^. The atypical category is especially problematic because many clinicians interpret it as “effectively negative”. TPS introduced diagnostic criteria designed to define “atypical urothelial cells” with emphasis on reporting major criterion of an NC ratio of at least 0.5, while relaxing assessment of other criteria (minor criteria): 1) nuclear hyperchromasia, 2) nuclear membrane irregularity and 3) chromatin quality. The criteria for atypia, in comparison to suspicious or positive, only requires identification of 2 (1 major, 1 minor criteria) of these features. Furthermore, atypia in TPS should exclude morphologic alterations attributable to a lithiasis or infections. Introduction of these diagnostic criteria under TPS serve to standardize assessment by encouraging assignment of a definitive diagnosis (negative, positive) or a slightly hedged diagnosis (suspicious) to avoid conflating loose criteria under PMC. However, reclassification under TPS has been shown to have relatively modest impact on the predictive value for UC despite avoidance of atypia classification, in part due to high interobserver variability and further complicated by the presence of other potential confounding benign cell types (e.g., seminal vesicle cells,, glandular cells, inflammatory cells, etc.). In addition, urothelial cell clusters can be challenging to assess due to ambiguous assessment criteria and blended cell borders.

Previous studies have sought to compare TPS to PMC to determine if the incidence of atypical diagnoses had changed using the standardized criteria ^17,18,20–27^. The prior studies have been cross-sectional in nature and were of limited sample size. Furthermore, few studies have assessed the impact of architectural barriers (e.g., excessive blood, confluent cellularity,) and human barriers (e.g., cytopathologist bias and human factors, i.e., fatigue, training, workload) for diagnosis ^28–31^.

In this study, we performed a longitudinal assessment of diagnostic differences between TPS and PMC in a retrospective cohort of nearly 1,300 specimens from 2008-2018, collected prior to the May 2018 implementation of TPS guidelines at an academic medical center, to assess how implementation of the TPS system influenced diagnoses. We hypothesized that, in agreement with previous studies, TPS assessments would yield fewer atypical assignments than PMC, but, uniquely, we were also interested in assessing and accounting for the time frame over which the adoption occurred. Longitudinal changes in PMC assessments may carry far-reaching implications for studies evaluating the implementation of cytology reporting systems, in that early, informal dissemination and discussion of materials might influence adoption. If such factors are identified, they could shed new light on optimal adoption strategies for new reporting criteria and technologies. A longitudinal assessment is important for determining the scope of such investigations.

## Methods

### Data Collection

After Institutional Review Board approval, we collected 1,277 urine samples across 141 patients (8 (Q1=5; Q3=12; IQR=7) repeat measurements per patient based on longitudinal follow-up) prior to incorporation of the TPS system at our institution, Dartmouth-Hitchcock Medical Center (DHMC). Sample collection did not reflect true incidence of UC classifications in the population, but were chosen to ensure a mix of different classifications for comparison between the two assessment methods. Urine samples, most of them voided, were prepared using ThinPrep® and Pap-stained prior to visual microscopic examination of the cytology slide. Pathologic examination was done using Olympus microscopes of various makes. Primary UC diagnoses were assigned at the time of collection by 10 different cytopathologists under the PMC system. The PMC system consisted of four diagnostic options (“Negative”, “Atypical”, “Suspicious”, and “Malignant”) without standardized criteria being applied.

After collecting PMC assessments from the original cytology report, all specimens were re-assessed by five cytopathologists explicitly considering TPS criteria; cytopathologists were not aware of the original diagnosis. Formal implementation of TPS occurred at DHMC in May 2018 and included the creation of specific TPS diagnostic categories within our Cerner Millenium system (“Negative for High Grade Urothelial Carcinoma”, “Atypical Urothelial Cells”, “Suspicious for High Grade Urothelial Carcinoma”, and “Positive for High Grade Urothelial Carcinoma”). Immediately prior to implementation, the cytopathologists met as a group to review TPS criteria, and the TPS criteria were presented at a genitourinary tumor board.

Four of the five pathologists who reassessed with TPS had also made previous assessments under PMC, but slides were selected to ensure the same pathologist did not assess the same slide twice. We also recorded whether the slide contained any significant cytological artifacts/confounders (e.g., blood, neobladder, confluent cellularity, etc.) that could impact assignment of high/low risk. Nondiagnostic and undetermined samples were removed from the analysis. We additionally recorded demographic information and collection date (**Table 1**). We also collected all PubMed articles directly referencing TPS across the study period by querying PubMed with the search term “The Paris System” and removing articles unrelated to urine cytology. For each article, we recorded the publication year and number of articles citing these publications by the writing of this work as a measure of lasting influence.

**Table 1:** Demographic characteristics of study participants; specimen source, history and whether an artifact (e.g., blood, confluent cellularity, neobladders and viewing artifacts, anything that could impact assessment quality) was present was registered on the specimen level, while patient demographics (age, sex) were registered on the patient level; for these tables information that could not be determined was removed

| Source | N (%) | History | N (%) |
| --- | --- | --- | --- |
| Barbotage | 6 (0.6) | Bladder Cancer | 943 (86.6) |
| Catheterized | 10 (0.9) | Hematuria | 110 (10.1) |
| Cystoscopic | 36 (3.3) | Other | 36 (3.3) |
| Ileal | 18 (1.7) |  |  |
| Neobladder | 7 (0.6) | Artifact Present N (%) | 236 (21.7) |
| Nephrostomy | 2 (0.2) |  |  |
| Voided | 986 (90.5) | Patient Demographics |  |
| Wash | 14 (1.3) | Age (mean (SD)) | 71.87 (10.94) |
| Other | 10 (0.9) | Sex = M (%) | 106 (75.7) |

### Diagnostic Differences between PMC and TPS

Assignment of UC categories with PMC and TPS were compared by cross-tabulating diagnoses into a table form to visualize changes between the two systems. Several Bayesian hierarchical logistic regression models were fit to the data in order to ascertain: 1) were cytopathologists operating under TPS less likely to assign atypical class, and 2) whether the PMC assignments began to resemble TPS nearing the end of the collection period (Appendix “Estimating Diagnostic Changes between PMC and TPS across Time”). Separately, overall agreement between the two systems was approximated through calculation of the intra-class correlation coefficient (*ICC*; Appendix “Overall Differences between PMC and TPS”). Pathologists who assessed fewer than ten slides were removed from the statistical analyses. Collection dates were mean-centered and units were scaled to years. Odds ratios (OR) were reported to communicate effect size. Statistical significance was assessed using calculation of the 95% high density posterior credible interval (CI), analogous to the confidence interval, and the probability of direction, *pd*, related to the p-value (*p* ≈ 2 ∗ (1 − *pd*); “p-value”). An OR and corresponding CI less than one indicates a negative association with the outcome or decreased likelihood, while greater than one indicates a positive association or increased likelihood given one unit increase in the predictor. Model fitting was accomplished using the R v3.6 statistical language via the *brms* package, with post hoc comparisons via the *emmeans* package ^32–36^.

## Results

### Comparing UC Assignments between TPS and PMC

Diagnoses (645 negative, 340 atypical, 83 suspicious, and 34 positive) were assigned by 10 different pathologists under the PMC system. Five pathologists provided UC assignments (722 negative, 254 atypical, 83 suspicious, and 43 positive) on the same set of cases from 2008-2018 using the TPS system (**Table 2**).

**Table 2:** Cross-tabulation of UC class assignments by cytopathologists operating under PMC and TPS across the entire study period (2008-2018)

|  | UC Class | TPS |  |  |  | Total |
| --- | --- | --- | --- | --- | --- | --- |
|  |  | Negative | Atypical | Suspicious | Positive |  |
| PMC | Negative | 601 | 44 | 0 | 0 | 645 |
|  | Atypical | 120 | 183 | 33 | 4 | 340 |
|  | Suspicious | 1 | 24 | 40 | 18 | 83 |
|  | Positive | 0 | 3 | 10 | 21 | 34 |
|  | Total | 722 | 254 | 83 | 43 | 1102 |

The cross-tabulated cytological findings in Table 2 suggest moderately high overall agreement between PMC and TPS (*ICC*=0.907, 95% CI: 0.883-0.929). Pathologists operating under PMC and TPS tended to agree on negative (*ICC=*0.908) and positive assignments (*ICC=*0.903), while generally in lesser agreement over assignments of atypical (*ICC=*0.744) and suspicious (*ICC=*0.775) UC classes (**Table 3**).

**Table 3:** Estimated intra-class correlation coefficients for each UC class as a measure of agreement between PMC/TPS

| UC Class | ICC | 2.5% CI | 97.5% CI |
| --- | --- | --- | --- |
| Negative | 0.908 | 0.876 | 0.937 |
| Atypical | 0.744 | 0.671 | 0.804 |
| Suspicious | 0.775 | 0.689 | 0.852 |
| Positive | 0.903 | 0.834 | 0.957 |
| Overall | 0.907 | 0.883 | 0.929 |

Notably, there were a few important findings that drove disagreement between the two systems. There were significantly fewer atypical assignments in TPS (n=254) as compared to PMC (n=340) (OR=0.37, 95% CI: [0.25-0.54], p<0.001; **Table 4**). A majority of the atypical cases that were reassigned to other UC classes under TPS system were downgraded to negative (n=120), compared to PMC (n=44) (OR=0.25, 95% CI: [0.15-0.42], p<0.001; **Table 4**), and as compared to other UC classes (suspicious, positive), which were reassigned to less frequently. A few atypical findings were reassigned to suspicious (n=33), though reassignments were marginally insignificant (OR=0.66, 95% CI: [0.40-1.04], p=0.07; **Table 4**).

**Table 4:**
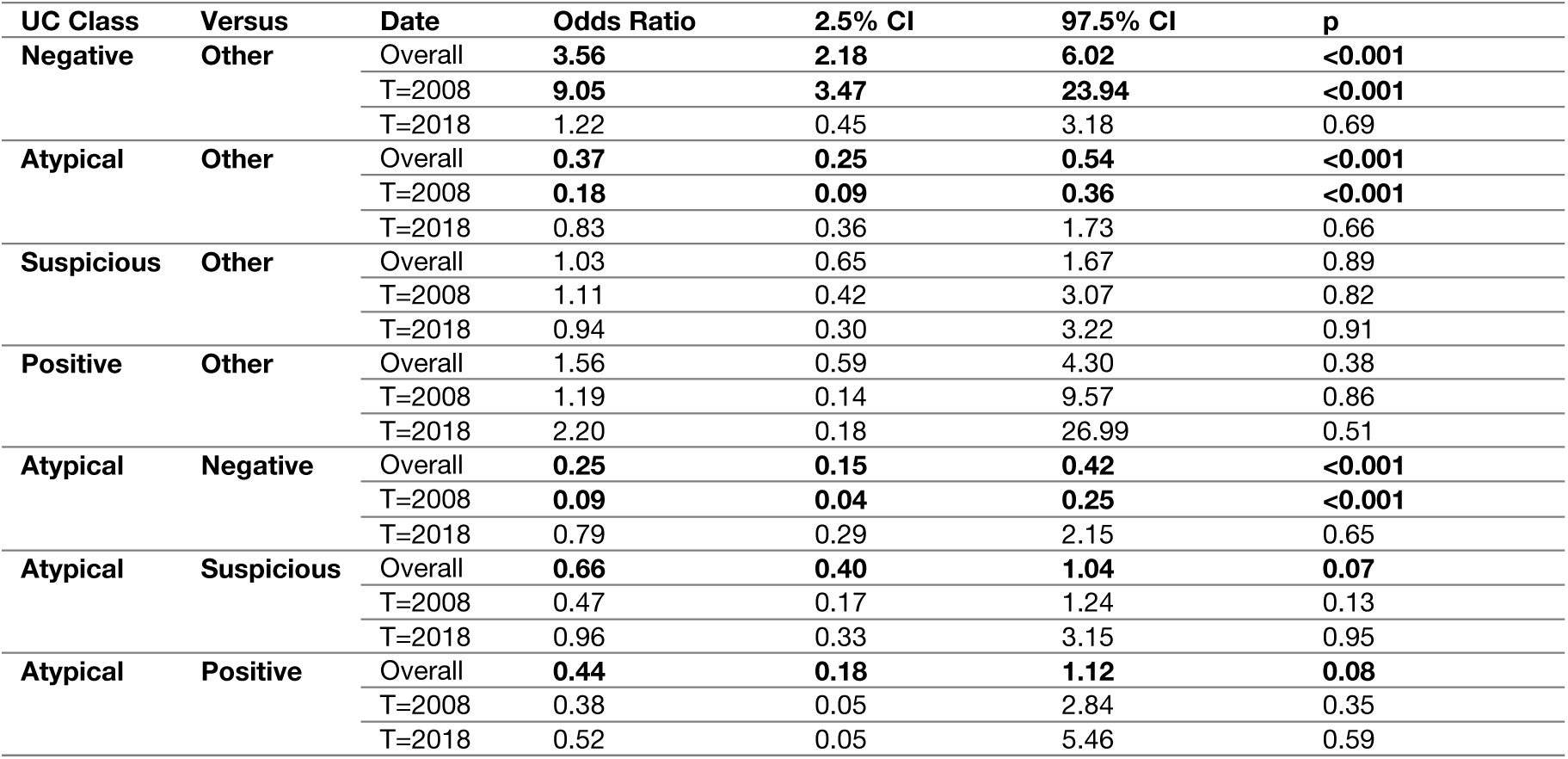
Reported effect estimates and “p-values” which compare odds of obtaining UC class given TPS versus odds of obtaining UC class given PMC; odds ratio greater than one indicates that assignment of UC was associated with TPS over PMC, whereas odds ratio less than one indicates UC assignment was associated with PMC over TPS; odds ratios were calculated: a) across all timepoints (*Overall*), and separately using the conditional effects from the interaction term– b) first date of collection in 2008 (*T=2008*) and c) last date of collection in 2018 prior to implementation of TPS (*T=2018*)

Interestingly, 71 specimens were reassigned from other UC classes under PMC to atypical under TPS. There were more negative and positive findings with TPS (n=765=722+43) as compared to PMC (n=679=645+34), while the number of suspicious findings did not change although 43 specimens deemed suspicious under PMC were reassigned under TPS (**Table 2**).

There appeared to be no impact of specimen artifacts on grading (Supplementary Table 1).

### Longitudinal Changes in UC Diagnostic Findings under PMC

Interestingly, assignment patterns of UC atypical and negative categories under PMC began to resemble TPS by the end of the collection period and the formal implementation of TPS. At the beginning of the study period, 2008, pathologists operating under TPS were far less likely to assign an atypical category as compared to PMC (OR=0.18, 95% CI: [0.09-0.36], p<0.001; **Table 4**), even more so than reported overall across the collection period (OR=0.37, 95% CI: [0.25-0.54], p<0.001; **Table 4**). However, at the end of the collection period, 2018, the differences between PMC and TPS for atypical assignments had entirely disappeared (OR=0.83, 95% CI: [0.36-1.73], p=0.66; **Table 4**). Conversely, the opposite effect can be seen for the negative UC class– larger differences between PMC and TPS at the start of the collection period (OR=9.05, 95% CI: [3.47-23.94], p<0.001; **Table 4**), even more so than reported across the collection period (OR=3.56, 95% CI: [2.18-6.02], p<0.001; **Table 4**), and then disappearance of the association near the formal implementation of TPS (OR=1.22, 95% CI: [0.45-3.18], p=0.69; **Table 4**). It should be noted that the time of assessment via PMC and re-assessment via TPS for 2008 cases is separated by more than a decade (2008 versus 2019), while the assessment time difference has diminished by 2018, hence suggesting potential similarities in reporting of negative cases towards the conclusion of the collection period. These associations were driven by reductions in atypical and increases in negative assignments for PMC, relative to TPS, such that assignment patterns of atypical and negative categories were similar between the two systems by the end of the collection period (2008 OR=0.09, 95% CI: [0.04-0.25], p<0.001; 2018 OR=0.79, 95% CI: [0.29-2.15], p=0.65). Through longitudinal modeling, the differences between these two systems may have disappeared as soon as 2016, when TPS was first officially published (**Figure 1A**). These findings appear to coincide with the rise in influential TPS-related publications (**Figure 1B**).

**Figure 1:**
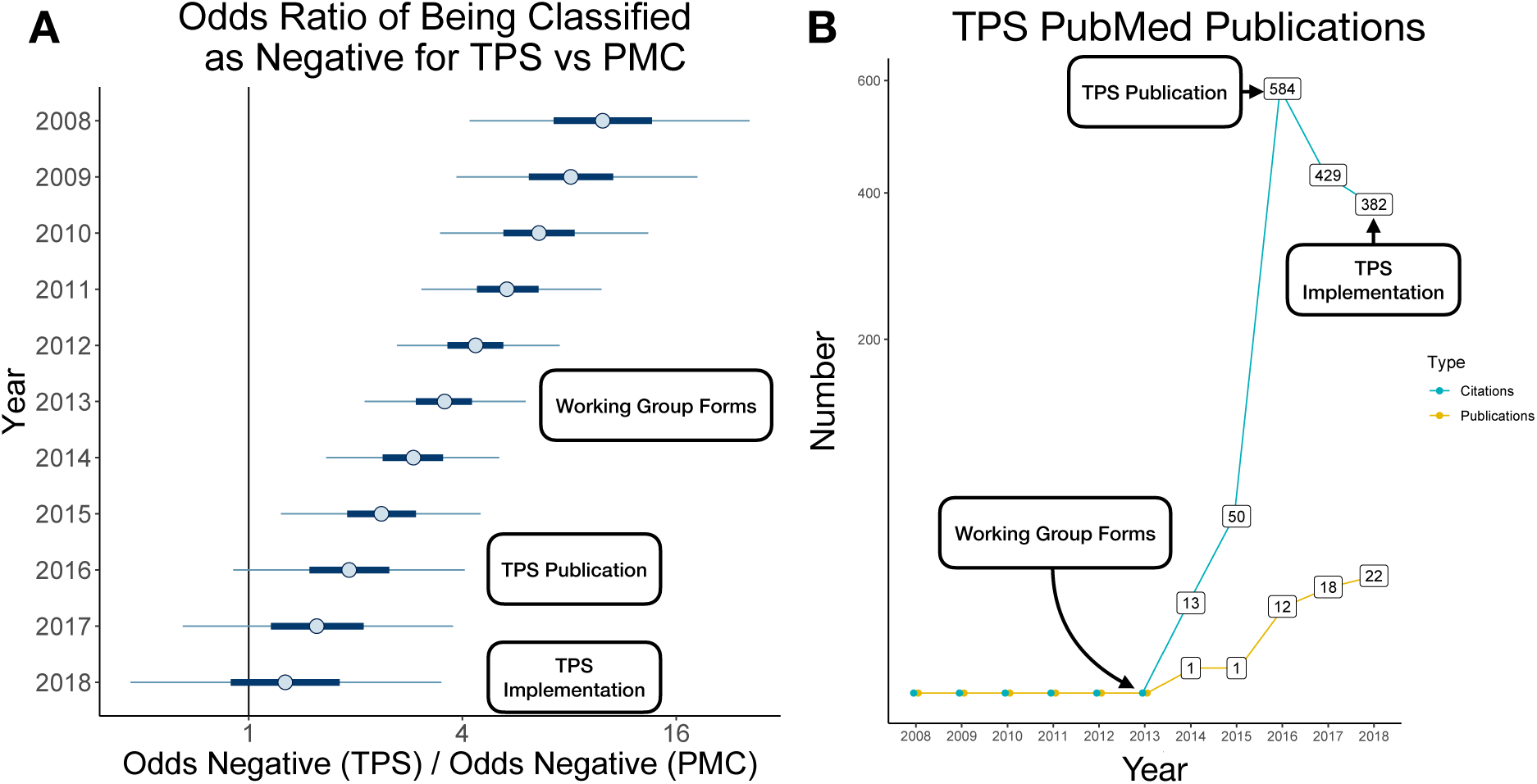
Longitudinal differences in negative UC class assignment under each system: **A)** Odds ratios (OR) and 95% CI (thin line) for assignment of negative UC class between TPS and PMC systems reported yearly in the month of May using the multinomial statistical model; OR of 1 (vertical line) indicates no difference in assignment of negatives between TPS and PMC; OR greater than one indicates that, for the same cases, more negative diagnoses were assigned using TPS as compared to PMC, with number of atypical UC diagnoses assigned as a reference (proxy for how PMC atypical diagnoses were reassigned as negative under TPS); results indicate that differences between the two systems disappeared by 2016, where the credible interval intersects OR=1; **B)** Number of yearly TPS-related publications across time and number of articles citing these publications by the writing of this article in 2022 as a proxy for article influence

## Discussion

Assessment of urine cytology is currently used for prospective screening and longitudinal monitoring, while cystoscopy and histological examination of a tissue biopsy provide the definitive diagnoses for urothelial carcinoma and dictate oncology care ^37–39^. Thus, a hybrid system has evolved in which cytology serves to triage patients to more or less invasive follow up procedures in a paradigm similar to cervical cancer screening. Reporting standardization enabled through adoption of well-defined quantitative and qualitative criteria (e.g., TPS) may soon make cytological analysis a viable alternative to a significant subset of cystoscopies and biopsies by reducing equivocal diagnoses or reports of “atypia” ^42^. This study sought to compare urine cytology assessment findings between the prior mode of cytologic assessment (PMC) and the Paris system (TPS) as well as to understand how urine screening assessment changed over the prior decade given informal adoption of standard TPS criteria.

Findings from this study corroborate previously published evidence that diagnoses under TPS discourage “atypia” classifications. In our study, we found that a vast majority of atypical cases under PMC were instead reclassified as negative under TPS. Many PMC atypical specimens were also relabeled as suspicious and positive, which suggests that the more definitive TPS criteria may have led to fewer equivocal diagnoses. However, these differences were not uniform over time, as differences between the PMC and TPS systems were greatest at the beginning of the study period but negligible immediately before formal implementation of TPS. This suggests several conclusions: 1) given that TPS was first discussed in 2013 and officially published in 2016, it is possible that cytopathologists may have been influenced by the discussion and publication of the guidelines, even though official adoption of the guidelines at our institution did not occur until mid 2018 (e.g., **Figure 1B** uses yearly publications and downstream citations to discuss the burgeoning influence of the guideline’s discussions and publication), and 2) previous studies failed to include longitudinal findings for the PMC/TPS comparison. By failing to stratify or adjust for modification across time, prior studies may have underestimated the degree to which these two systems initially diverged, as the atypical category was assigned in far greater numbers by PMC as compared to reassessment by TPS for 2008. The gap between PMC and TPS had disappeared before 2018, just prior to the implementation of TPS. These findings have potential implications across other pathology subdisciplines, where timely adoption of new criteria is of interest as evolving staging/grading guidelines feature more quantitative endpoints. Efforts to establish rigorous quantitative endpoints for disease pathology may be further accelerated using computer-assisted diagnostic decision aids (e.g., machine learning) ^1,43,44^.

Because several large studies have already examined the differences between TPS and PMC in their ability to predict definitive diagnoses from cystoscopy and/or biopsy, we opted to not assess concordance of TPS/PMC with these independent measures. Neither did we seek to compare differences in assignment patterns between and “within” pathologists via estimates of inter-and intra-observer reliability, which has been well characterized in the literature and could have potentially impacted final effect estimates (e.g., changes in clinical staff). We did not assess changes in diagnostic assignment per pathologists across time (i.e., years of experience, training), which presents an interesting follow-up direction. Other limitations include differences between real clinical sign-out (as was seen with PMC) and academic/research sign out (seen with TPS) (e.g., TPS raters did not have access to clinical information or the cytotechnologist’s preliminary diagnoses, both of which can influence final diagnosis in practice). The lack of clinical information may hamper the ability to apply TPS criteria for the final UC assessment, as cytologists regularly look for and consider factors such as stones that can cause reactive atypia. While associations were found that were longitudinal in nature, we are unable to definitely conclude that publication of TPS influenced assessment prior to adoption of the system in our institution due to the potential presence of temporal unmeasured confounders. Some potential unmeasured confounders are known (e.g., attendance at conferences, interactions with other practicing cytologists, professional social media exchanges as means to get information–e.g., Twitter, #CytoChat) while others may be unknown and unanticipated. Nonetheless, this study evaluated agreement amongst two cytology assessment systems longitudinally over a range not previously covered by any similar study.

The study findings point to the potential role and impact of widespread discussions and communication on early adoption, dissemination, and implementation of evolving clinical guidelines. While TPS is now commonplace, reasons for longitudinal differences in PMC prior to TPS include early publications, presentation of TPS criteria at conferences, interactions and communication among pathologists across various clinical and virtual settings, and differences in training/experience. However, reasons for change are nearly impossible to evaluate as such data is not routinely collected, which precludes the application of statistical methodologies (e.g., social network analysis, difference-in-differences design, causal inference) that could elucidate such findings. As similar cytology reporting systems are rolled out or refined (e.g., The Bethesda systems for thyroid and cervical cytopathology, Milan system for salivary gland cytopathology, and other standardized reporting systems, such as fluid cytology, breast cytology, etc.), there exist opportunities to develop and apply innovative data collection and analytical methods to build a greater understanding of successful guideline adoption strategies. At a minimum, retrospective comparisons to previous assessment methods prior to official adoption of new criteria should be performed over a broad time horizon (e.g., 4-5 years as suggested by the significant study findings) sufficient to characterize changing practices.

## Conclusion

TPS purports to improve reporting of UC by defining clear criteria for diagnostic categories. In our retrospective study, we provide confirming evidence of the achievement of intended changes in diagnostic assignment. A unique longitudinal assessment suggested that early adoption of standard reporting criteria may have occurred prior to the implementation of TPS in our institution. This will hardly be the last change in reporting practices as increasingly quantitative and reproducible reporting criteria are adopted. As these practices shift towards quantitative endpoints, expert time and specialization required for nuanced decision making may increase, though partial automation of expert decision making may ameliorate its most challenging aspects. In the future, development and adoption of semi-autonomous classification systems may present the next step forward in the development of standard reporting criterion, as these computerized systems can augment the laborious process of cytological inspection and potentially pave the way for adoption of urine cytology as an alternative for cystoscopy for high-grade urothelial carcinoma.

## Data Availability

The datasets presented in this article are not readily available because of participant privacy concerns. Requests to access the datasets should be directed to.

