## Appendix for "Large-Scale Longitudinal Comparison of Urine Cytological Classification Systems Reveals Potential Early Adoption of The Paris System Criteria"

#### Estimating Diagnostic Changes between PMC and TPS across Time

Several binomial, multinomial and ordinal logistic regression models were fit to the data to estimate diagnostic differences between PMC and TPS. Repeat measurements within patient, specimen and cytopathologist were accounted for with mixed effects modeling and potential bias was accounted for using Bayesian modeling.

First, we related diagnostic assignment to whether the specimen was assessed via PMC or TPS via the following statistical model:

$$y_i \sim Binomial(1, p_i)$$
$$logit(p_i) = \beta_0 + \beta_1 I(\text{TPS}_i) + \beta_2 \text{artifact}_i + \beta_3 \text{time}_i + \beta_4 I(\text{TPS}_i) * \text{time}_i + \theta_{\text{rater}[i]} + \theta_{\text{patient}[i]} + \theta_{\text{specimen}[i]}$$
$$\theta_{\text{rater}[i]} \sim N(0, \tau^2_{\text{rater}})$$
$$\theta_{\text{patient}[i]} \sim N\left(0, \tau^2_{\text{patient}}\right)$$
$$\theta_{\text{specimen}[i]} \sim N\left(0, \tau^2_{\text{specimen}}\right)$$
$$\beta \sim MVN(0, v\mathbf{I})$$

The aforementioned model compares one UC class to all of the other classes (e.g., atypia versus negative, suspicious, and positive; i.e., to find whether atypia assignments changed between the two systems). We also fit multinomial models to perform pairwise comparisons between each of the UC classes (e.g., negative versus atypical) to corroborate with findings from the cross-tabulated table (i.e., were some of the atypical assignments reassigned to suspicious under the new system?), where the outcome is represented as: $y_i \sim Categorical(p)$. Atypical was used as a reference group for comparison to the other UC classes. Fixed effects were included that: 1) assessed changes under TPS as compared to PMC ($I(\text{TPS}_i)$), 2) how assignments changed over collection dates ($\text{time}_i$), and 3) whether assignments changed over time for PMC to assess whether there was some evidence of informal adoption of TPS prior to implementation of TPS via the interaction $I(\text{TPS}_i) * \text{time}_i$. Interaction findings were communicated through report of the conditional effects of TPS/PMC at the start of the collection period (2008) to the end of the collection period (2018). Separately, we removed the interaction term to assess for overall TPS/PMC changes, independent of collection date. These statistical models also controlled for cytological artifacts/confounders (e.g., blood, neobladder, confluent cellularity, scanning artifacts) that may have impacted diagnosis ($\text{artifact}_i$) and variation within and between clusters of repeat measurements ($\theta_{\text{rater}[i]}$, $\theta_{\text{patient}[i]}$, $\theta_{\text{specimen}[i]}$).

#### Overall Differences between PMC and TPS

Overall concordance between the two measurement systems was measured using the intra-class correlation coefficient, *ICC*, which is related to the weighted Kappa measure used for assessing agreement in measurements. Since UC classes are ordered by risk (negative, atypical, suspicious, positive), we calculated the *ICC* using an ordinal regression model, of the same functional form as the binomial and multinomial models with the following changes:

$$y_i \sim Categorical(\text{p})$$
$$p = [p_1 = q_1, p_2 = q_2 - q_1, p_3 = q_3 - q_2, p_4 = 1 - q_3]$$
$$logit(q_j) = \kappa_j - \mu_i$$
$$\mu_i = \beta_0 + \beta_1 I(\text{TPS}_i) + \beta_2 \text{artifact}_i + \beta_3 \text{time}_i + \beta_4 I(\text{TPS}_i) * \text{time}_i + \theta_{\text{rater}[i]} + \theta_{\text{patient}[i]} + \theta_{\text{specimen}[i]}$$
$$\kappa_j \sim N(0, v_2)$$

Where it is assumed that some continuous "marker" of disease severity underlies the UC classes (although assignment of "atypia" is potentially unclear in this context) and that binning this distribution with cutpoints κ is the cytopathologist's attempt to approximate the true disease severity with a UC class. The *ICC* is given by:

$$ICC = \frac{\tau_{rater}^2 + \tau_{patient}^2 + \tau_{specimen}^2}{\tau_{rater}^2 + \tau_{patient}^2 + \tau_{specimen}^2 + \frac{\pi^2}{3}}$$

We also calculated *ICC* measures for the binomial and multimodal models where appropriate and also reported *ICC* measures by year through addition of year-specific specimen random intercepts to the aforementioned statistical models. Increases in *ICC* by year would provide further evidence that PMC more closely resembles TPS prior to the implementation of TPS.

### Supplementary Tables

**Supplementary Table 1:** Impact of artifacts on UC class calls

| UC Class | Odds Ratio | 2.5% CI | 97.5% CI | p |
|---|---|---|---|---|
| **Negative** | 1.52 | 0.59 | 3.70 | 0.37 |
| **Atypical** | 0.84 | 0.48 | 1.56 | 0.60 |
| **Suspicious** | 1.12 | 0.50 | 2.54 | 0.78 |
| **Positive** | 0.27 | 0.04 | 1.49 | 0.13 |
| **Overall** | 0.56 | 0.24 | 1.34 | 0.19 |
